# Effects of Randomized Corruption Audits on Early-Life Mortality in Brazil

**DOI:** 10.1101/2020.09.14.20194597

**Authors:** Simeon Nichter, Antonio P. Ramos, Leiwen Gao, Robert E. Weiss

## Abstract

**Background:** Various studies suggest that corruption affects public health systems across the world. However, the extant literature lacks causal evidence about whether anti-corruption interventions can improve health outcomes. We examine the impact of randomized anti-corruption audits on early-life mortality in Brazil.

**Methods:** The Brazilian government conducted audits in 1,949 randomly selected municipalities between 2003 and 2015. To identify the causal effect of anti-corruption audits on early-life mortality, we analyse data on health outcomes from individual-level vital statistics (*DATASUS*) collected by Brazil’s government before and after the random audits. Data on the audit intervention are from the *Controladoria-Geral da Uniao*, the government agency responsible for the anti-corruption audits. Outcomes are neonatal mortality, infant mortality, child mortality, preterm births, and prenatal visits. Analyses examine aggregate effects for each outcome, as well as effects by race, cause of death, and years since the intervention.

**Results:** Anti-corruption audits significantly decreased early-life mortality in Brazil. Expressed in relative terms, audits reduced neonatal mortality by 6.7% (95% CI −8.3%, −5.0%), reduced infant mortality by 7.3% (−8.6%, −5.9%), and reduced child mortality by 7.3% (−8.5%, −6.0%). This reduction in early mortality was higher for nonwhite Brazilians, who face significant health disparities. Effects are greater when we look at deaths from preventable causes, and show temporal persistence with large effects even a decade after audits. In addition, analyses show that the intervention led to a 12.1% (−13.4%, −10.6%) reduction in women receiving no prenatal care, as well as a 7.4% (−9.4%, −5.5%) reduction in preterm births; these effects are likewise higher for nonwhites and are persistent over time. All effects are robust to various alternative specifications.

**Interpretation:** Governments have the potential to improve health outcomes through anti-corruption interventions. Such interventions can reduce early-life mortality and mitigate health disparities. The impact of anti-corruption audits should be investigated in other countries, and further research should further explore the mechanisms by which combating corruption affects the health sector.

## 1 Introduction

Although early-life mortality has fallen substantially across the world in recent decades, it remains disturbingly high in many countries. Moreover, significant disparities in child survival rates persist across ethnicity and socioeconomic characteristics in much of the world.^1^ In part to reduce early-life mortality, the WHO and other international organizations have embraced Universal Health Coverage (UHC), which was adopted as a Sustainable Development Goal by all UN member states. Achieving UHC will require substantial investments, estimated to be an additional $370 billion per year across low and middle-income countries.^2^ In addition to this expanded coverage, research suggests that millions of deaths across all ages could be prevented if additional resources improved low-quality healthcare.^3^

Despite these potential benefits, increased healthcare expenditures do not always improve health outcomes, especially in countries with poor governance.^4^ Among various reasons is corruption — the abuse of public office for private gain — a phenomenon that siphons off significant healthcare investments in much of the world. According to Transparency International (2019), corruption in the healthcare sector leads to world-wide losses of over $500 billion per year, more than the amount necessary to provide Universal Health Coverage across the globe.^5^ For example, public officials in many contexts pilfer funds earmarked for medical equipment and exchange bribes for suboptimal healthcare contracts. Evidence suggests that corruption has substantial effects not only on healthcare expenditures, but also on health outcomes. For example, Hanf et al. (2016) estimate that corruption in the healthcare sector kills approximately 140,000 children across the world each year.^6^

For decades, researchers and practitioners largely ignored the pernicious effects of corruption on health. By contrast, publications in leading medical journals over the past few years have called for increased attention to the “hidden pandemic” of corruption.^6–8^ Yet many unanswered questions remain about how to address this pressing problem. Various interventions have been proposed to combat corruption in the healthcare sector, such as improving top-down controls, heightening bottom-up accountability, and raising salaries.^5^ And numerous case studies and analyses of observational data investigate the effectiveness of specific measures, usually with mixed findings. But a systematic investigation by the Cochran Review (2016) found no published studies providing causal evidence about whether anti-corruption interventions can affect health outcomes.^9^

To address this lacuna, the present article provides novel evidence that one such intervention — randomized anti-corruption audits — improved early-life mortality in Brazil. We identify the causal impact of these audits on neonatal mortality, infant mortality and child mortality. To explore mechanisms, we estimate the effects of audits on prematurity — a leading cause of early-life mortality — and on prenatal visits, which can reduce these deaths.^10^ Given substantial disparities in healthcare, we also examine effects across race; information on race is included in vital statistics, and is highly associated with socioeconomic disparities in Brazil. In addition, analyses examine effects by cause of death and years since the intervention. Before turning to statistical analyses, we first provide context about public healthcare in Brazil, especially about to the role of municipalities and how audits can affect health outcomes.

## 2 Background on Healthcare and Corruption in Brazil

Brazil’s 1988 Constitution declares the universal right to comprehensive healthcare. The public healthcare system, known as *SUS* (*Sistema Único de Saúde*, or Unified Health System) serves the substantial majority of Brazilians, as only a quarter of the population enrolls in private health plans.^11^ Although Brazil’s expenditures on healthcare grew nearly fivefold between 2000 and 2012, *SUS* faces numerous challenges including understaffing of doctors, lengthy delays for specialized services, and limited medical supplies.^12^ Indeed, healthcare is frequently identified as the most pressing problem in Brazilian public opinion surveys, with 87% of respondents in a recent survey rating the quality of health clinics and hospitals as “low” or “very low.”^13^ While various health indicators have improved markedly in recent years, substantial regional and socioeconomic inequities in health outcomes persist.^14–16^ For example, infant mortality fell from 77 to 14 deaths per thousand births between 1980 and 2016, but infants born in the relatively poor Northeast states of Bahia and Piauí have double the mortality rates as those born in wealthier southern states of Rio Grande do Sul and Santa Catarina.^17,18^ Given that *SUS* is the primary provider of healthcare in Brazil, most neonatal, infant and child deaths occur within the public healthcare system — motivating our investigation of whether political corruption is a cause of early-life mortality in Brazil.

As one of the most decentralized countries in the world,^19^ Brazil places healthcare responsibilities on all levels of government, including its 5,570 municipalities that are governed by elected mayors who wield considerable power. These municipalities implement primary care and many other aspects of healthcare, using funds primarily from the federal government. Federal transfers to municipalities account for nearly 48 percent of all public health expenditures, and municipal officials enjoy substantial autonomy and minimal oversight when expending much of these funds.^11,20^ For example, municipalities receive significant resources to distribute free medicine through Brazil’s Popular Pharmacy Program *(Programa Farmácia Popular do Brasil*), but municipalities often do not follow top-down procurement guidelines, and federal audits reveal poor monitoring of the inflow and outflow of medications from municipal clinics and hospitals.^20^ While civil society may provide oversight through municipal health councils, these councils often have limited autonomy, particularly in smaller municipalities where they depend on funds from local governments to operate.^21^

Corruption is a pressing concern in contemporary Brazil.^22–24^ Over the past few years, scores of politicians and bureaucrats have been prosecuted as part of the nation’s largest-ever corruption investigation (*Operação Lava Jato*, or Operation Car Wash),^25,26^ and President Jair Bolsonaro railed against corruption as a pillar of his campaign platform.^27,28^ Corruption often involves healthcare. For example, the “Blood-suckers” (*Sanguessugas*) corruption scandal — involving extraordinarily overpriced ambulances — implicated nearly a hundred politicians who reportedly received kickbacks and enabled these wasteful purchases.^29,30^ During the current COVID-19 crisis, which has caused over 125,000 Brazilian deaths, corruption probes have implicated several state governors and dozens of public officials for similar infractions involving the procurement of medical supplies.^31^ Moreover, in a major cross-national survey, 10.9% of all Brazilian respondents reported paying a bribe to access healthcare during the past year.^32^ Many policymakers suggest a link between corruption and poor health outcomes. For instance, a justice on Brazil’s Supreme Court recently emphasized: “Corruption kills. It kills in the waiting line of *SUS* [the public healthcare system], in the lack of hospital beds, in the lack of medicine.”^33^

To combat corruption in Brazil’s federal expenditures, the *Controladoria-Geral da União* (*CGU*, or Office of the Comptroller General) was formed in 2003. That year, the *CGU* initiated an impressive audit program to root out the corrupt use of federal funds by municipalities. This program, entitled *Programa de Fiscalizaçāo por Sorteios Públicos* (Monitoring Program with Public Lotteries), randomly selected municipalities in televised lotteries every few months, and sent teams of auditors for extensive visits to scrutinize their expenditures of federal funds. By early 2015, the *CGU* had conducted 40 lotteries and performed audits in 1,949 municipalities, involving a careful investigation of over R$22 billion of federal funds.^34^ Municipalities were randomly selected for audits in 40 lotteries, with stratification at the state level. The lotteries excluded municipalities with populations above 500,000, which were ineligible for the intervention; less than one percent of Brazilian municipalities exceed this threshold.

The *CGU* audits reveal substantial corruption in the sphere of healthcare. For example, the audit of Capelinha in Minas Gerais discovered that the municipality’s financial records included many false receipts for medicine that was in fact never purchased for its public health clinics.^35^ Avis et al. (2018) show that the *CGU*’s audits substantially reduce the level of corruption for years after a municipality is audited.^34^ One important reason for this subsequent decline in corruption is that when these audits uncover corruption, politicians face electoral as well as legal punishments.^36^ We leverage random assignment of these anti-corruption audits to identify their causal effect on early-life mortality.

## 3 Methods

### 3.1 Data Sources and Definitions

We use health outcomes from individual-level vital statistics collected by the Brazilian government. Data on deaths is taken from the *Sistema de Informação sobre Mortalidade* (*SIM*) database, while births is taken from the *Sistema de Informações sobre Nascidos Vivos (SINASC*) database. Both databases are maintained by Ministry of Health’s Information Technology division *(DATASUS*) and include individual-level details about all births and all deaths across the nation. For each year from 2004 to 2016, we obtained information on births and deaths. These figures reflect all Brazilian municipalities with fewer than 500,000 inhabitants. Information about preterm births and prenatal visits are also from *SINASC*. Data on the dates of audits are from the *CGU*, the government agency responsible for the anti-corruption audits. For each year from 2003 to 2015, these data indicate whether a municipality was selected by lottery for a corruption audit. Municipal populations was obtained from the Brazilian Institute of Geography and Statistics (IBGE).

The primary outcomes are counts of deaths, stratified by age group (neonatal, infant, and child), cause of death (all causes and preventable causes) and race (white and nonwhite). Neonatal mortality refers to deaths before 28 days of age, infant mortality refers to deaths before one year of age, and child mortality refers to deaths before five years of age. Our database includes information on 24,088,335 births and 463,611 child, 394,860 infant deaths and 270,087 neonatal deaths.

Preventable causes follow standard definitions from the International Classification of Diseases (ICD-10). The two racial groups are based on the five categories provided in the *SINASC* and *SIM* databases. In addition to using the “white” (*branca*) category, we aggregate the other four categories as “nonwhite”: “black” (*preta*), “brown” (*parda*), “Asian” (*amarela*), and “indigenous” (*indígena*).

Additional outcomes include prenatal visits and preterm births. For prenatal visits, the *SINASC* dataset provides three categories: 0, 1-6, and 7+ visits with healthcare professionals. Preterm births refer to births before 37 gestational weeks. Both variables are also stratified by race (white and nonwhite). Tables 1-3 provide information about each outcome variable.

**Table 1.** Deaths by Race and Age Group (2004-16)

|  | All | White | Nonwhite | Missing Race |
| --- | --- | --- | --- | --- |
| Neonatal Deaths | 270,087 | 105,605 | 125,816 | 38,666 |
| Infant Deaths | 394,860 | 158,157 | 184,357 | 52,346 |
| Child Deaths | 463,611 | 185,362 | 220,161 | 58,088 |

*Note:* Data reflect all Brazilian municipalities with under 500,000 inhabitants, as municipalities exceeding this threshold are ineligible for the *CGU*'s randomized audits. Neonatal deaths are less than 28 days of age. Infant deaths are cumulative deaths less than one year of age. Child deaths are cumulative deaths less than five years of age.

**Table 2.** All Births and Preterm Births, by Race (2004-16)

|  | All Races | White | Nonwhite | Missing Race |
| --- | --- | --- | --- | --- |
| All Births | 24,088,335 | 10,162,130 | 13,110,423 | 815,782 |
| Preterm Births | 190,225 | 78,895 | 87,457 | 23,873 |

*Note:* Data reflect all Brazilian municipalities with under 500,000 inhabitants, as municipalities exceeding this threshold are ineligible for the *CGU's* randomized audits. Preterm births refer to births before 37 gestational weeks.

**Table 3.** Number of Prenatal Visits per Birth, by Race (2004-16)

|  | All Races | White | Nonwhite | Missing Race |
| --- | --- | --- | --- | --- |
| 0 Visits | 550,643 | 119,160 | 393,473 | 38,010 |
| 1-6 Visits | 9,688,857 | 2,864,615 | 6,421,463 | 402,779 |
| 7+ Visits | 13,804,811 | 7,109,314 | 6,175,695 | 519,802 |

*Note:* Data exclude municipalities with over 500,000 inhabitants, which are ineligible for the *CGU's* randomized audits. The *SINASC* dataset provides three categories: 0, 1-6, and 7+ visits by healthcare professionals. The second column, from top to bottom, shows the number of births from any race that had received 0, 1-6, and 7+ prenatal visits (respectively). The next columns shows analogous data for births that were white, nonwhite, and with missing race data (respectively).

### 3.2 Model

We model death as Bayesian hierarchical Poisson model, as deaths are count data with a highly skewed, nonnegative distribution. Our data is indexed by municipality *i* = 1,… *N* and year *j* = 1,… *J*. Outcomes *y_ij_* are counts of deaths for municipality *i* in year *j*. Municipality effects are modelled as random intercepts *β_i_*; year effects *γ_j_* are modelled as having a priori an AR(1) autocorrelation structure.

In our main specification, audit effects are modelled as an indicator variable *A_ij_* in municipality *i* and year *j*. For the municipalities that has been audited *A_ij_* =0 for all years up to the year of the audit and *A_ij_* = 1 afterward. If the municipality has never been audit *A_ij_* = 0 for all years. The parameter *μ_ij_* is the expected number of deaths in municipality *i* in the year *j*. The offset is log(*b_ij_*), the number of births in municipality *i* in year *j*. Our full model is

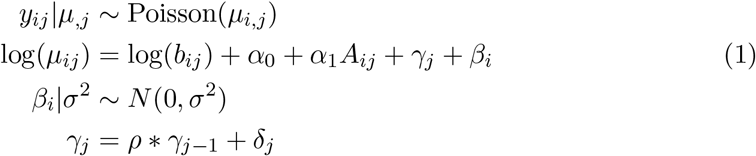

for *j* > 1, and

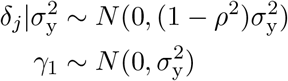

where *α*_0_ is the intercept, *α*_1_ is the audit effect, and *ρ* is the correlation between year effects. Municipality random effects *β_i_* model unmeasured, time-invariant differences in the baseline condition among municipalities. These municipality random effects also account for some overdispersion. The *γ_j_* year effect models common national dynamics in the decline of mortality and follows an autoregressive process. Bayesian methods are employed to fit the models; priors are weakly informative.

### 3.3 Additional Specifications

Whereas Model 1 above estimate the effects of anti-corruption audits on outcomes is all years following the audit year, another specification investigates the extent to which these effects vary and persist over time. Model 2 is identical to Model 1, except the treatment variable is partitioned into 10 effects one for each of the next 10 years after an audit: *ψ_k_* for each year *k* = 1,… 10. In this specification, the term *α*_1_*A_ij_* in Equation (1) is replaced by 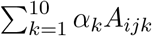. To examine heterogeneity of audit effects with respect to race (whites and nonwhites) and cause of death (preventable and all causes), we fit Models 1 and 2 using subsets of the data.

### 3.4 Inferential Targets

Across all specifications, the key quantity of interest is the coefficient for audit effects, either *A_ij_* or 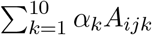. This quantity can be interpreted as the *relative mortality reduction* caused by anti-corruption audits, comparing the treatment group against the control group. Using model 1, we also estimate the number of deaths averted in treated municipalities, from one year after their respective audits until 2016.

### 3.5 Robustness

Numerous analyses were employed to ensure robustness of findings to modelling and data choices. We also fit Poisson models and negative binomial models using maximum likelihood estimation, with fixed effects for municipalities and years. In addition, we used alternative approaches to deal with the fact that some municipality-years have no births to be included as offsets. While our preferred specification addresses this issue by using births+1 as the offset, we also confirmed robustness to alternative specifications. Additional information about these robustness checks is included in the appendix.

## 4 Results

**Figure 1 shows trends in neonatal, infant and child mortality between 2004 and 2016 in Brazil, based on vital statistics data employed in this study**. For every thousand births, neonatal mortality declined from 13 in 2004 to 10 in 2016, infant mortality declined from 20 to 14, and child mortality declined from 23 to 17.

**Figure 1.**
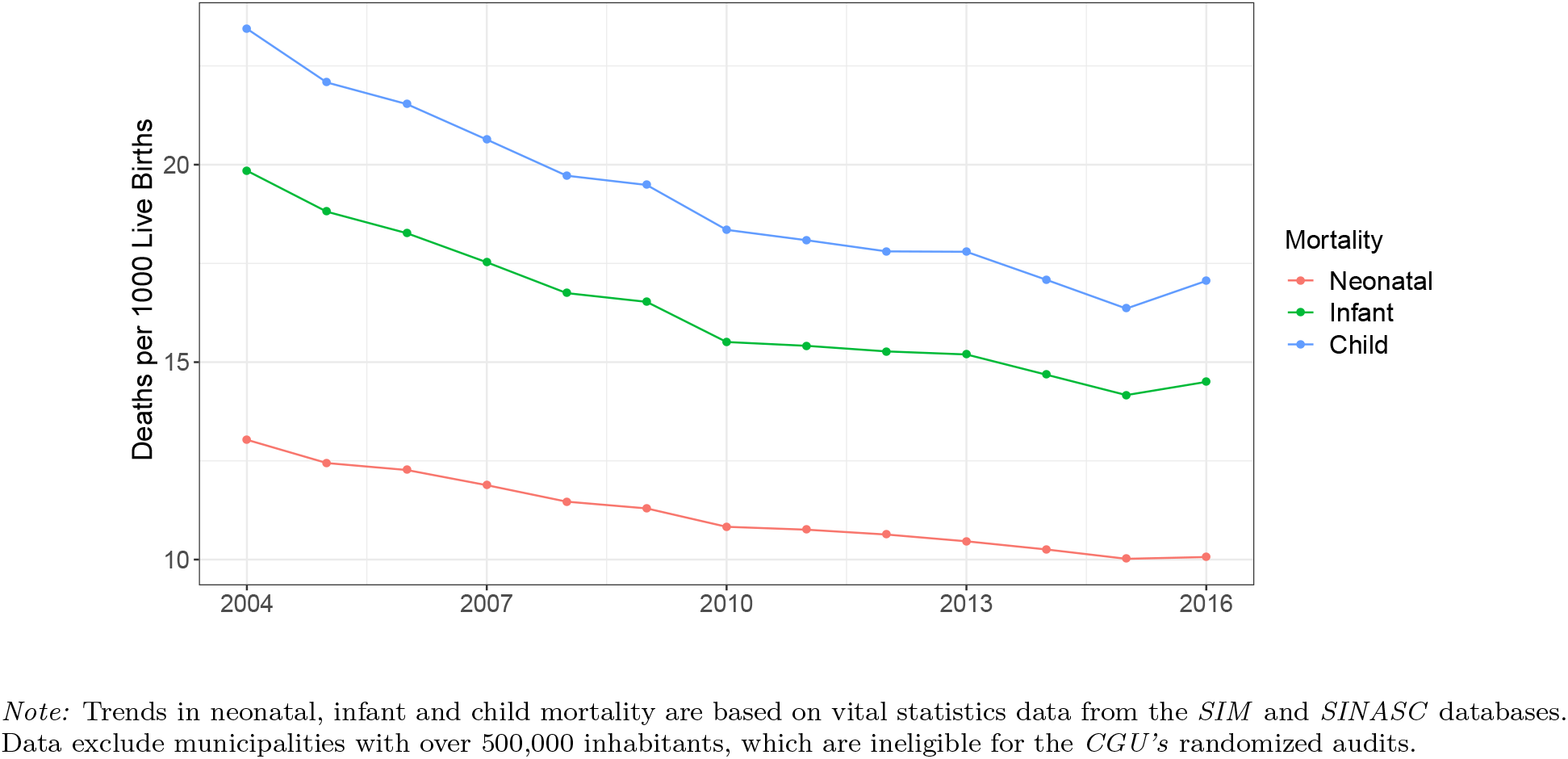
Trends in Early-Life Mortality in Brazil (2004-2016)

**Figure 2 shows the effect of anti-corruption audits on early-life mortality in Brazil**. The left panel reports results from Model 1 fitted with data for all causes of death. Dots represent point estimates and error bars reflect 95% credible intervals. The first specification fits the model using data for all races. Anti-corruption audits reduced neonatal mortality by 6.7% (95% CI −8.3%, −5.0%), reduced infant mortality by 7.3% (−8.6%, −5.9%), and reduced child mortality by 7.3% (−8.5%, −6.0%). The next two specifications fit the model using data for white and nonwhite Brazilians, respectively. Audits decreased neonatal mortality among whites by 3.3% (−6.1%, −0.5%), decreased infant mortality by 3.6% (−6.0%, −1.3%), and decreased child mortality by 4.0% (−6.1%, −1.8%). Among nonwhites, audits reduced neonatal mortality by 8.1% (−10.3%, −5.7%), reduced infant mortality by 8.2% (−10.1%, −6.3%), and reduced child mortality by 8.2% (−9.9%, −6.4%).

**Figure 2.**
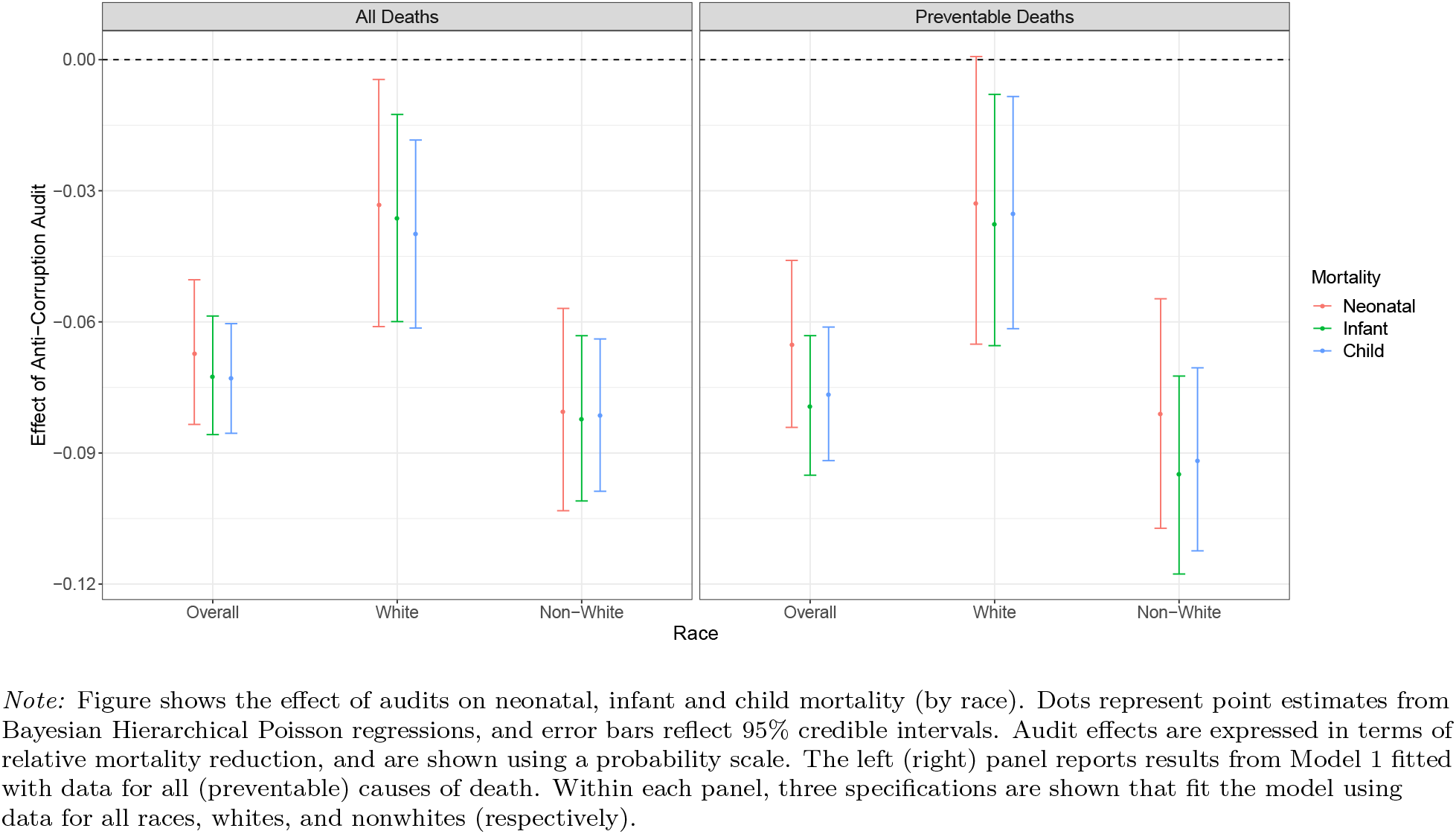
Effect of Anti-Corruption Audits on Early-Life Mortality.

Findings are similar for preventable deaths. The right panel of Figure 2 analogously shows results from Model 1, fitted only with data for deaths by preventable causes. Across all races, audits reduced neonatal mortality by 6.5% (95% CI −8.4%, −4.6%), reduced infant mortality by 7.9% (−9.5%, −6.3%), and reduced child mortality by 7.7% (−9.2%, −6.1%). Again, this reduction in early-life mortality is smaller among white than nonwhite Brazilians. Audits decreased neonatal mortality among whites by 3.3% (−6.5%, 0.1%), decreased infant mortality by 3.8% (−6.5%, −0.8%), and decreased child mortality by 3.5% (−6.2%, −0.8%). By contrast, audits reduced neonatal mortality among nonwhites by 8.1% (−10.7%, −5.5%), reduced infant mortality by 9.5% (−11.8%, − 7.2%), and reduced child mortality by −9.2% (−11.2%, −7.1%). All other results discussed below reflect deaths from all causes, but are robust to focusing on preventable causes only. hs only.

**Figure 3 reports results from Model 2, in which treatment effects of the intervention may vary over time**. The left panel fits the model using data for all races. After one year, audits reduced neonatal mortality by 1.7% (95% CI −4.1%, 0.7%), reduced infant mortality by 2.6% (−4.5%, −0.6%), and reduced child mortality by 2.5% (−4.3%, −0.7%). While these effects were relatively small, and only statistically significant for infant and child mortality, audits’ impact increased with time. After five years, audits decreased neonatal mortality by 7.8% (−10.3%, −5.1%), decreased infant mortality by 8.3% (−10.4%, −6.1%), and decreased child mortality by 8.0% (−9.9%, −6.0%). And after ten years, audits reduced neonatal mortality by 13.3% (−16.6%, − 9.9%), reduced infant mortality by 15.0% (−17.7%, −12.3%), and reduced child mortality by 14.1% (−16.6%, −11.6%).

**Figure 3.**
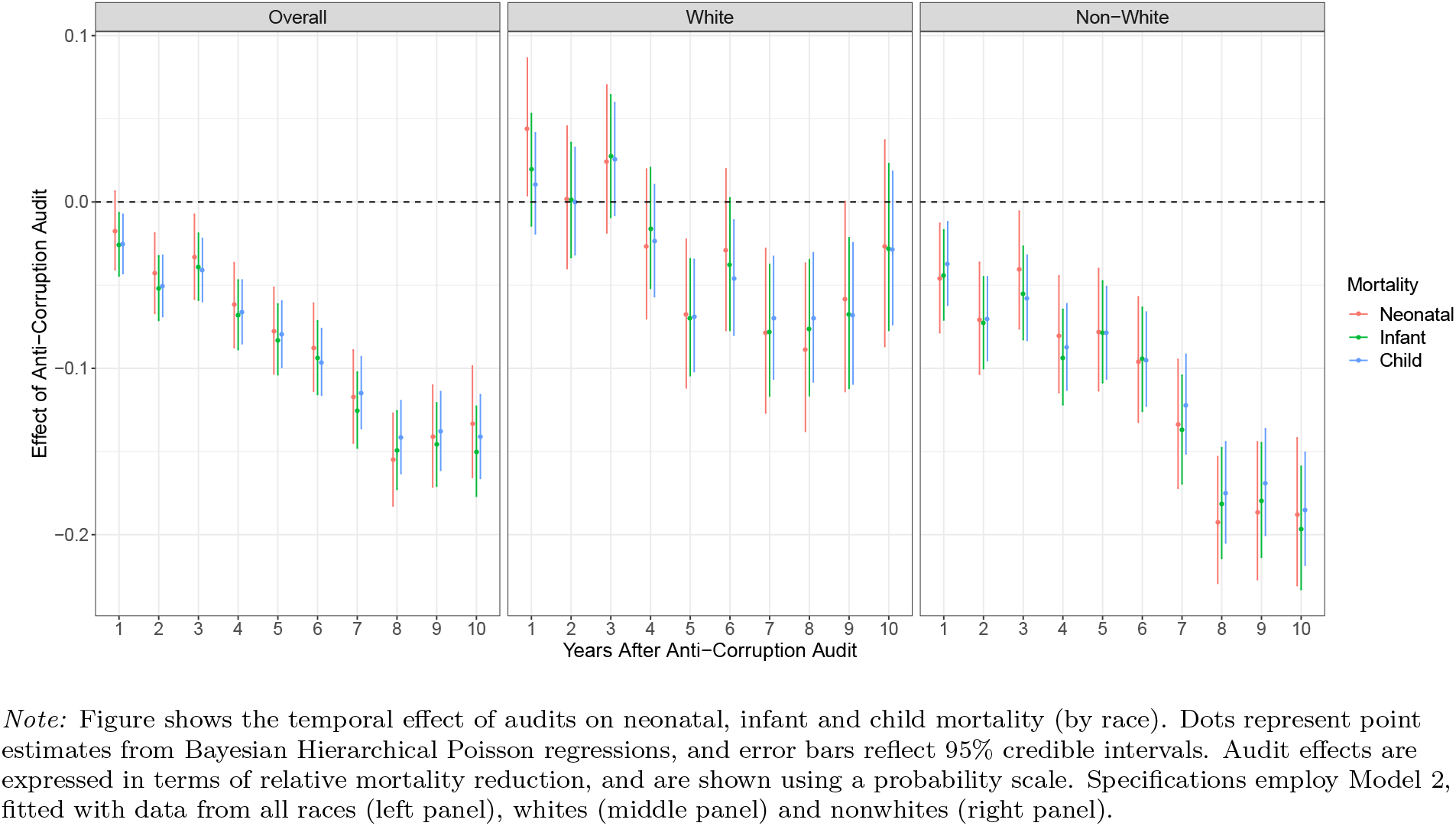
Temporal Effects of Anti-Corruption Audits on Early-Life Mortality.

The next two panels of Figure 3 stratify deaths by race. For whites, a significant reduction was only observed in three of ten years for neonatal mortality (5, 7 and 8 years post-audit), in four years for infant mortality (5, 7, 8 and 9 years post-audit), and in five years for child mortality (5-9 years post-audit); the magnitude of these significant effects ranged from −4.6% to −8.9% (middle panel). By contrast, the intervention significantly decreased all three mortality measures for nonwhites in all ten years after an audit (right panel). After one year for nonwhites, audits reduced neonatal mortality by 4.6% (95% CI −7.9%, −1.3%), reduced infant mortality by 4.4% (−7.1%, −1.7%), and reduced child mortality by 3.7% (−6.2%, −1.2%). After five years, audits decreased neonatal mortality in this population group by 7.8% (−11.4%, −4.0%), decreased infant mortality by 7.9% (−10.9%, −4.7%), and decreased child mortality by 7.9% (−10.6%, −5.1%). And after ten years for nonwhites, audits reduced neonatal mortality by 18.8% (−23.1%, −14.2%), reduced infant mortality by 19.7% (−23.3%, −15.9%), and reduced child mortality by 18.5% (−21.8%, −15.0%).

**Figure 4 presents effects of anti-corruption audits on prenatal visits and preterm births**. Results are from Model 1, with specifications fitting the data for all races, whites, and nonwhites. With respect to prenatal care, the intervention led to a 12.1% reduction (95% CI −13.4%, −10.6%) in women receiving no prenatal visits. This decrease was 14.3% (−15.8%, −12.9%) for nonwhite Brazilians and 4.0% (−6.9%, −1.1%) for white Brazilians. In addition, audits caused a 2.8% fall (−3.1%, −2.5%) in women receiving 1-6 prenatal visits. This effect was a 1.1% decrease for nonwhites (−1.5%, −0.8%), and insignificant for whites. By contrast, audits increased women receiving at least 7 prenatal visits by 4.8% overall (4.6%, 5.1%). This increase was 6.0% (5.6%, 6.4%) for nonwhites, but only 1.3% (0.9%, 1.6%) for whites.

**Figure 4.**
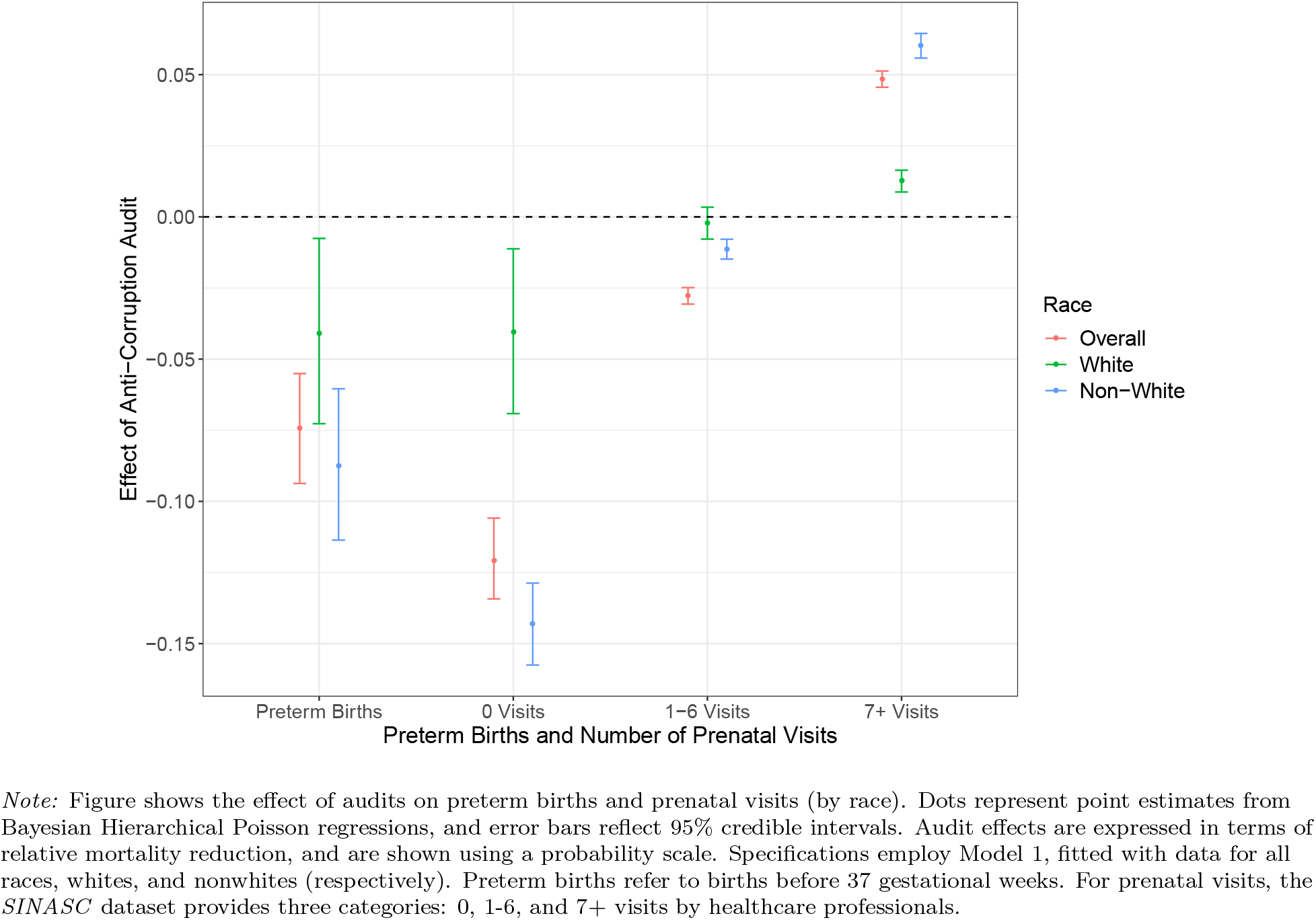
Effect of Anti-Corruption Audits on Preterm Births and Prenatal Visits.

Figure 4 shows that anti-corruption audits reduced births before 37 gestational weeks by 7.4% (95% CI −9.4%, −5.5%). This decrease was 8.8% (−11.4%, −6.0%) for nonwhite Brazilians, versus 4.1% (−7.3%, −0.8%) for white Brazilians. Next, Figure 5 reports results from Model 2, in which the effect of audits on prematurity may vary over time. The left panel fits the model using data for all races, and finds no effect in the first year after an audit, but significant negative effects in each subsequent year. For example, preterm births fell 6.5% (−9.6%, −3.4%) five years after an audit, and fell 16.2% (−19.9%, −12.3%) ten years after an audit. The next two panels of Figure 5 also reflect results from Model 2, but fit the data only for whites and nonwhites, respectively. For whites, a significant reduction in preterm births was observed for the 7th, 8th, 9th and 10th year after an audit; the point estimates on these effects range from −7.8% to −11.2% (middle panel). For nonwhites, no effect is observed in the first post-audit year, but significant negative effects are observed in each year thereafter (right panel). Audits decreased preterm births among nonwhites by 8.6% (−12.6%, −4.5%) after two years, by 5.7% (−10.2%, −1.2%) after five years, and by 21.3% (−26.5%, −16.0%) after ten years.

**Figure 5.**
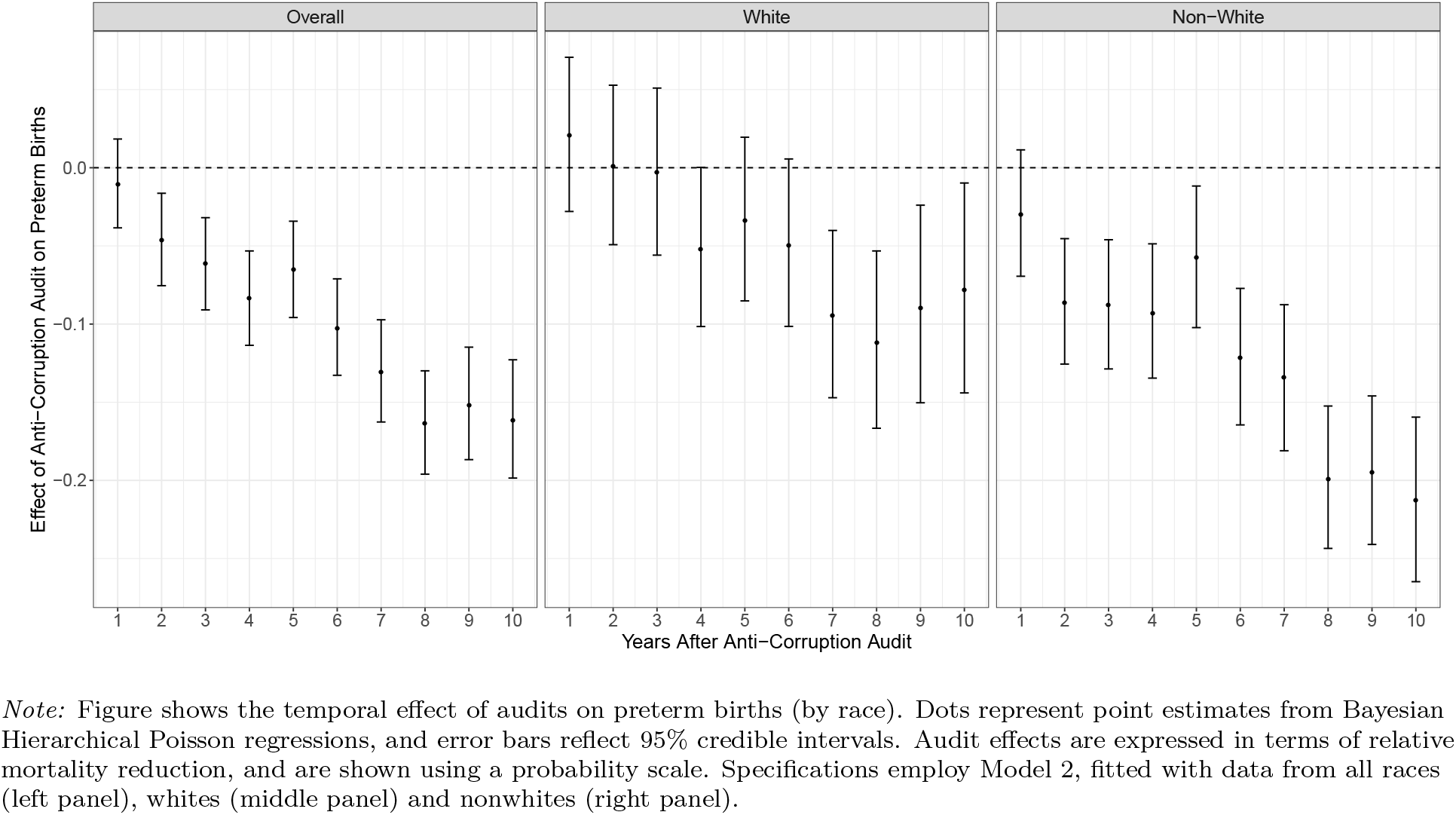
Temporal Effects of Anti-Corruption Audits on Preterm Births.

**Table 4 estimates the number of lives saved in audited municipalities as a result of the anti-corruption intervention**. Between 2004 and 2016, 129,783 children under the age of five perished in audited municipalities. Our results suggest that 139,985 deaths would have occurred without the intervention, suggesting that audits saved the lives of 10,202 children during this period. Nonwhite children represent over three-quarters of averted deaths.

**Table 4.** Lives Saved by Anti-Corruption Audits (2004-2016)

|  | *Actual Deaths* |  |  | *Counterfactual Deaths* |  |  | *Lives Saved* |  |  |
| --- | --- | --- | --- | --- | --- | --- | --- | --- | --- |
|  | All Races | Whites | Nonwhites | All Races | Whites | Nonwhites | All Races | Whites | Nonwhites |
| 2004 | 2,795 | 963 | 1,291 | 3,015 | 1,003 | 1,406 | 220 | 40 | 115 |
| 2005 | 6,543 | 2,311 | 2,918 | 7,057 | 2,407 | 3,177 | 514 | 96 | 259 |
| 2006 | 8,391 | 3,181 | 4,046 | 9,051 | 3,313 | 4,405 | 660 | 132 | 359 |
| 2007 | 9,164 | 3,466 | 4,516 | 9,884 | 3,610 | 4,917 | 720 | 144 | 401 |
| 2008 | 10,266 | 3,751 | 5,373 | 11,073 | 3,907 | 5,850 | 807 | 156 | 477 |
| 2009 | 10,485 | 3,723 | 5,675 | 11,309 | 3,878 | 6,179 | 824 | 155 | 504 |
| 2010 | 10,971 | 3,984 | 5,900 | 11,833 | 4,150 | 6,424 | 862 | 166 | 524 |
| 2011 | 11,543 | 4,278 | 6,049 | 12,450 | 4,456 | 6,586 | 907 | 178 | 537 |
| 2012 | 11,766 | 4,295 | 6,282 | 12,691 | 4,474 | 6,839 | 925 | 179 | 557 |
| 2013 | 12,171 | 4,376 | 6,585 | 13,128 | 4,558 | 7,169 | 957 | 182 | 584 |
| 2014 | 12,019 | 4,407 | 6,527 | 12,964 | 4,590 | 7,106 | 945 | 183 | 579 |
| 2015 | 11,844 | 4,348 | 6,521 | 12,775 | 4,529 | 7,100 | 931 | 181 | 579 |
| 2016 | 11,825 | 4,455 | 6,536 | 12,755 | 4,640 | 7,116 | 930 | 185 | 580 |
| Overall | 129,783 | 47,538 | 68,219 | 139,985 | 49,516 | 74,273 | 10,202 | 1,978 | 6,054 |

*Note:* Left panel shows actual deaths in audited municipalities, from one year after their respective audits until 2016. These data are from the *SIM* database. Middle panel uses Model 1 to estimate the counterfactual number of deaths in audited municipalities, from one year after their respective audits until 2016. In this counterfactual, the municipalities were not audited. In right panel, lives saved is the difference between actual deaths and counterfactual deaths. Within each panel, data are shown for all races, whites, and nonwhites. Deaths averted with missing race data are included in “All Races,” but not shown separately.

**Finally, Figure 6 shows the results of the temporal effects of the audit program on child mortality over time — using model 2**. Models was fitted use births from all races, whites and non-white. Left panel presents results for 0 visits, the middle panel for 1-6 visits and the right panel for 7+ visits. The audits largely reduced the number of births with zero visits and the effects increase over time and are similar for all races. The audits mostly reduce the number of births with 1-6 prenatal visits. Results tend to be statistically significant but substantively unimportant; results are similar for all races. The audits statistically and substantively increased the number of births with more than seven or more prenatal visits; results are larger for non-white births.

**Figure 6.**
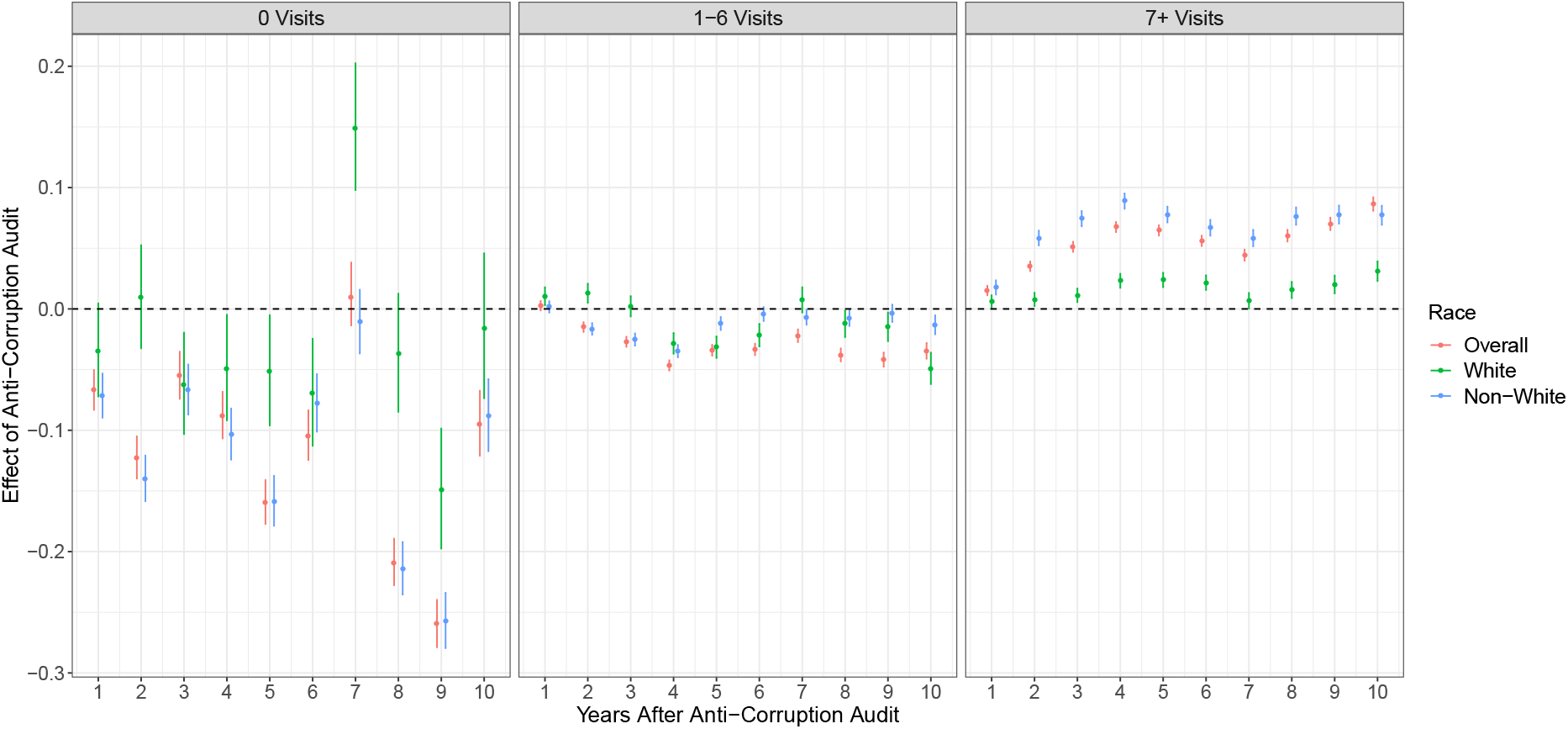
Temporal Effects of Anti-Corruption Audits on prenatal visitas. Coefficientes for the audit effects are plotted in probabilty scale. Results from model 2 fitted with data from all races (left panel), whites (middle panel) and non-white (right panel). Dots are points estimates error bars represent 95 % credible intervals.

## 5 Discussion

The findings of this study provide causal evidence that anti-corruption interventions can improve health outcomes. Between 2003 and 2015, the Brazilian government randomly selected 1,949 municipalities for extensive audits of how federal transfers were spent. Analyses suggest that these randomized anti-corruption audits decreased early-life mortality in Brazil. Expressed in relative terms, audits led to a 6.7% decline in neonatal mortality, a 7.3% decline in infant mortality, and a 7.3% decline in child mortality. The decrease in each type of mortality was at least twice as large for non-white Brazilians — who face significant health disparities — as for white Brazilians. The impact of audits on early-life mortality is greater when stratifying on deaths from preventable causes. Audits have a relatively small immediate effect, which magnify substantially over time and demonstrate temporal persistence. The intervention also improved prenatal care and reduced prematurity, which are commonly understood to be determinants diversion of early-life mortality.^10^ More specifically, audits caused a 12.1% decrease in women receiving no prenatal care, and 7.4% decrease in preterm births — findings that are likewise amplified for non-white Brazilians and persist over time. Overall, these results suggest that anti-corruption audits improved health outcomes.

The impact of audits was also substantial when considering the absolute number of lives saved by the intervention. Altogether, municipalities randomly selected for anti-corruption audits had 10,202 fewer child deaths in 2004-2016 than the counterfactual in which they had not been audited. This magnitude is considerable, given that 129,783 children under the age of five perished in these municipalities during this time period. The intervention disproportionately saved the lives of nonwhite Brazilians, who represent over three-quarters of averted deaths.

Several mechanisms may explain why randomized audits in Brazil reduced early-life mortality. One potential mechanism is that audits caused local public officials to pilfer less of the municipal budget, thereby reducing diversion of funds allocated to public healthcare. Avis et al. (2018) shows that corruption is 7.9% lower in municipalities that previously experienced random anti-corruption audits in Brazil, compared to unaudited municipalities.^34^ Less corruption would be expected to reduce diversion in health budgets, because many corrupt acts discovered by these audits involve officials diverting federal transfers earmarked for the public health system. For example, CGU audits have revealed municipal officials purchasing apartments with funds allocated to health projects, submitting fake receipts for medicine that was never purchased, over-invoicing medical purchases in exchange for kickbacks, and awarding bids to shill companies that never delivered public services.^35^ Reducing such diversion in health budgets increases funds available for various expenditures, some of which may reduce early-life mortality. For example, WHO (2018) identifies prematurity as the leading cause of death worldwide for children under five – and emphasizes that improved care can avert over three-fourths of fatalities of premature newborns.^10^ In addition to improved healthcare during and after childbirth, preterm births (and early-life mortality) can be reduced through prenatal visits with health professionals who play key roles in screening for risk factors, conducting ultrasounds, and providing information about dietary requirements and substance use.^10^ Given frequent disparities in access to these services in Brazil and beyond,^37–39^ increasing such expenditures may be especially likely to improve health outcomes of underrepresented groups. Although not dispositive, our results are consistent with this mechanism in which audits reduce the diversion of healthcare funds that can directly or indirectly reduce the deaths of young Brazilians: we find that anti-corruption audits increased prenatal visits, decreased preterm births, and reduced early-life mortality.

Another potential mechanism is that anti-corruption audits increased the quality of municipal politicians, who in turn engaged in less corruption and adopted policies reducing early-life mortality. Although democratic elections enable voters to punish corrupt politicians, citizens often lack information about the extent to which their elected representatives abuse public office for private gain.^36,40^ Whereas the mechanism discussed above reduces corruption by existing officials, the *CGU*’s widely disseminated audit results may also have alleviated this information asymmetry and allowed voters to punish local corrupt politicians at the ballot box. Consistent with this possibility, Ferraz & Finan (2008) show that Brazil’s randomized audits decreased the reelection of mayors – the chief executives of municipal governments – in municipalities with above-average levels of corruption.^36^ In contexts where corrupt mayors had diverted resources from the health sector, electing challengers may lead to reduced diversion from health budgets. Furthermore, challengers may also adopt policies that reduce early-life mortality, either by directly channeling resources to the public health system or through indirect channels such as infrastructure investments in water and sanitation. Beyond ousting corrupt incumbents, audits may also increase the quality of municipal politicians by improving the pool of challenger candidates. Many local politicians have been prosecuted using information gleaned from audits, which may dissuade corrupt individuals from entering politics if they update their perceptions of how risky it is to obtain corrupt rents as an elected official.^34^ More broadly, such political effects may help to explain the results of our study, which does not explore this possible mechanism.

An important avenue for future research is thoroughly investigating potential mechanisms underlying the findings of this study. With regards to the first mechanism discussed above, one next step would be to examine whether the anti-corruption audits increased expenditures on public goods – both in the public health system and more broadly – that are generally understood to improve early-life mortality. Existing studies do not definitely address this question: Avis et al. (2018) indicates that audits did not affect overall health spending,^34^ while an unpublished study by Lichand et al. (2017) finds that audits decreased spending of federal transfers earmarked for health.^411^ Another fruitful direction is to elaborate and test possible political mechanisms through which audits reduce early-life mortality. Such work should carefully consider reasons why voters may have supported corrupt politicians in the past: whereas many scholars posit the informational argument presented above, others suggest that voters may also deliberately choose corrupt politicians who effectively provide public goods or other benefits.^40^ Furthermore, this line of investigation should examine whether audits affect the creation of municipal councils, which promote civil society participation in health policy and have been shown to reduce infant mortality in Brazil.^43^ Overall, elaborating and unpacking such theoretical mechanisms can shed light on why anticorruption audits reduce early-life mortality in the Brazilian context.

While our results provide causal evidence that anti-corruption interventions can reduce early-life mortality, another key direction for future research is examining broader implications of the present study. With respect to generalizability, it is important to clarify whether government audits reduce deaths in many countries, and to understand scope conditions if the relationship is rarely or never observed elsewhere. Our findings also underscore the importance of examining the potential health impacts of a wider array of anti-corruption interventions, including other ways of heightening top-down controls as well as bottom-up accountability. Despite growing attention to effects of corruption on health, our findings suggest that increased attention to such questions can potentially save many lives, especially among vulnerable populations.

## Data Availability

The data is available from the mentioned sources in the paper.

## Appendix A: Distribution of Outcome Variables

**Figure 7.**
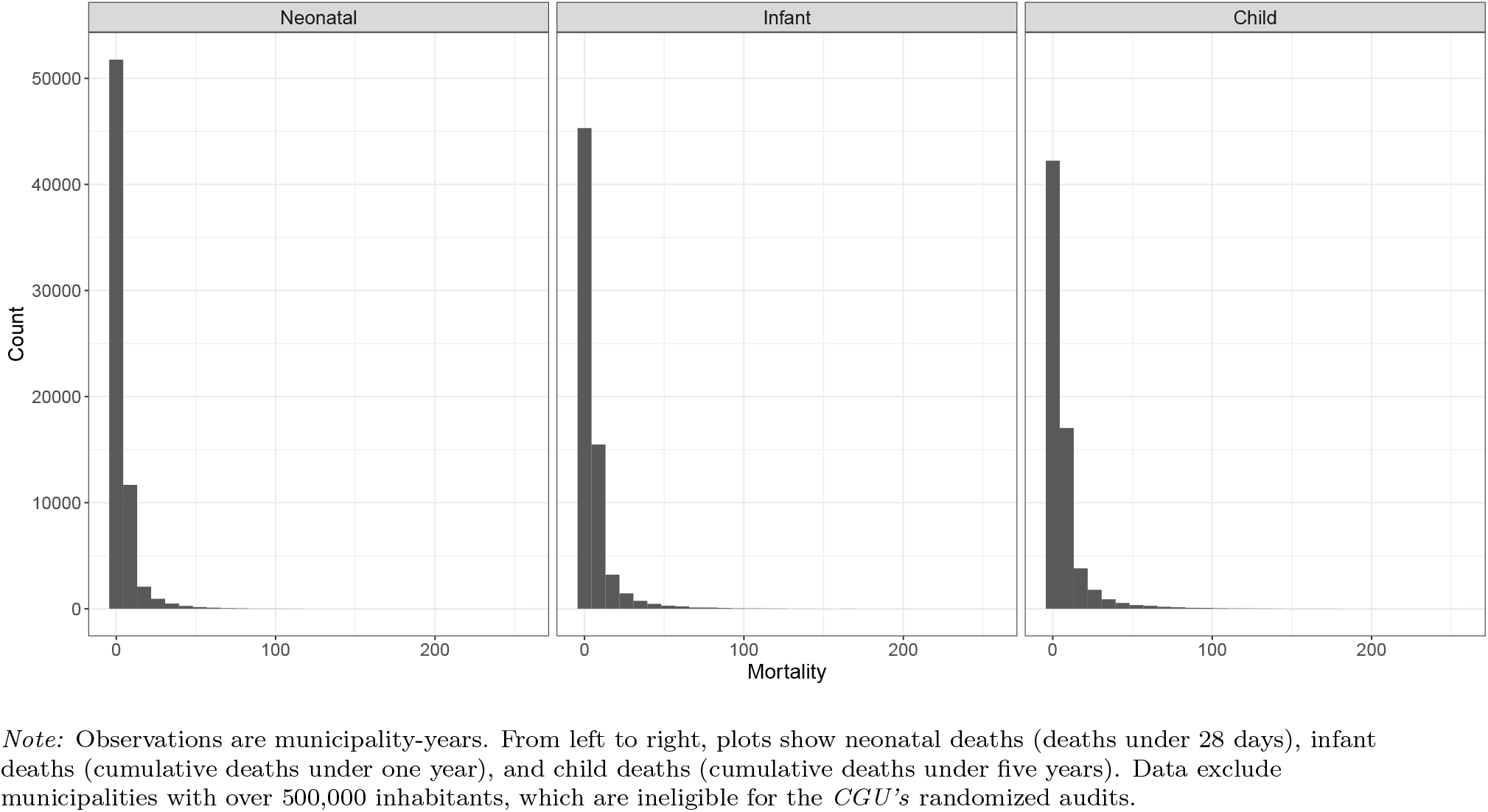
Distribution of Deaths (Municipality-Years, 2004-2016)

## Appendix B: Model Comparisons

We investigate the robustness of results to modeling and estimation procedures. Figure 8 shows a comparison of point estimates and confidence intervals for: (a) Poisson regression with fixed effects for municipalities and years estimated using maximum likelihood (ML); (b) negative binomial models with fixed effects for municipalities and years estimated using ML; and (c) the Bayesian hierarchical Poisson model with an AR(1) autocorrelation structure used in Models 1 and 2. Results are shown for neonatal mortality (left), infant mortality (middle), and child mortality (right). Across Figure 8, results are very similar with respect to the point estimates. Negative binomial model show large intervals for the point estimates, which is sensible as negative binomial models have an additional parameter to be estimated compared to the Poisson model – the over-dispersion parameter. This exercise shows that our point estimates are robust to estimation procedure (maximum likelihood or Bayesian), as well as fixed versus random effects.

**Figure 8.**
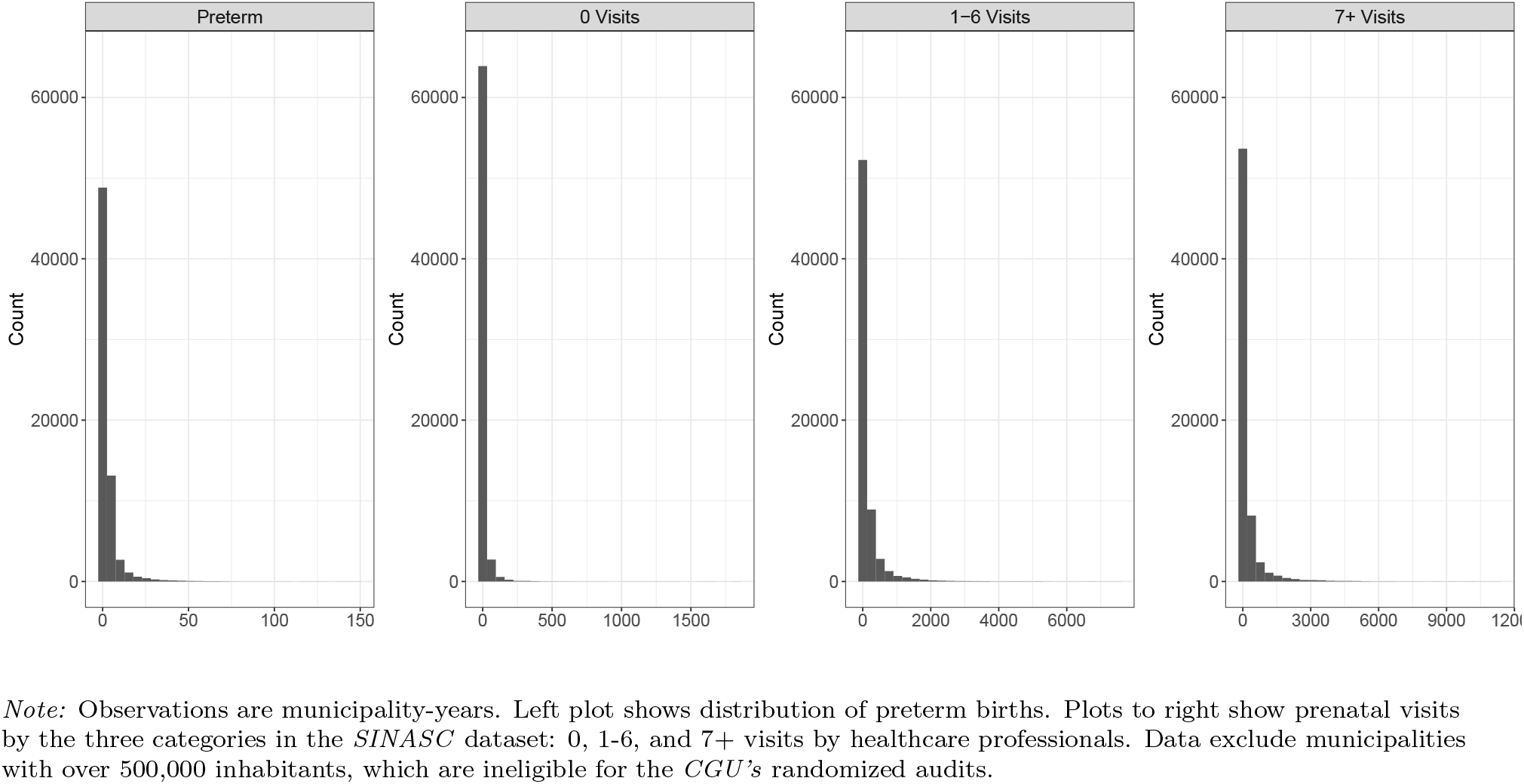
Distribution of Preterm Births and Prenatal Visits (Municipality-Years, 2004-2016)

## Appendix C: Offset Choice for Poisson Models

While using poisson regression to model mortality rates we need information on both the outcome variable, which is the deaths count and an offset parameter, which is the birth count. In the equation 1 they are, respectively, *y_i_*, *j* and *b_i_*,*j*. However, for some municipalities there is no birth and/or death information in specific years. Table ?? shows the percentage of municipality-year with 0’s births and/or deaths for mortality. The second column is the percentage of municipality-year with 0 birth as well as 0 death, while the third column incudes the percentage of municipality-year only with 0 birth but the count of death is not 0. Overall, by summing the last two columns up, there are at around 22.9% municipality-year with 0 birth. To deal with this issue, we apply three methods to manipulate the data in municipality-year level:

- Method 1: Add one birth to births counts across all observations.
- Method 2: Exclude every observation with 0 birth and add 1 to the count of births for all remaining observations.
- Method 3: Exclude only observations with 0 birth and 0 deaths and add 1 to the count of births for all remaining observations.

Figure 10 exhibits estimates for the effect of anti-corruption audits on three types of mortality, by cause of death using three methods. Since the estimates are similar using three methods, we choose method 1 to manipulate datasets in all models, which includes the largest number of observations without losing any information.

**Figure 9.**
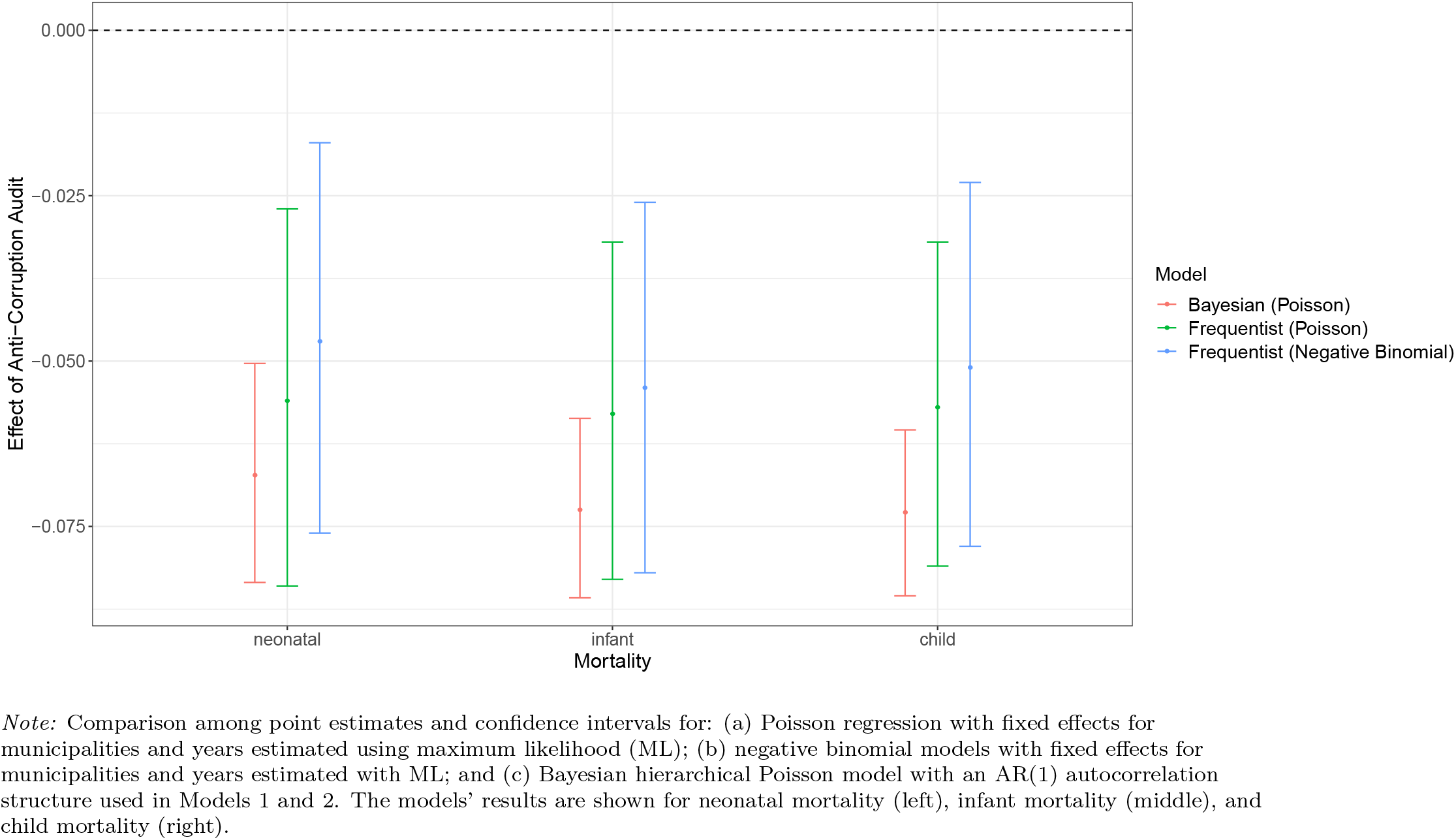
Model Comparisons.

**Table 5.**
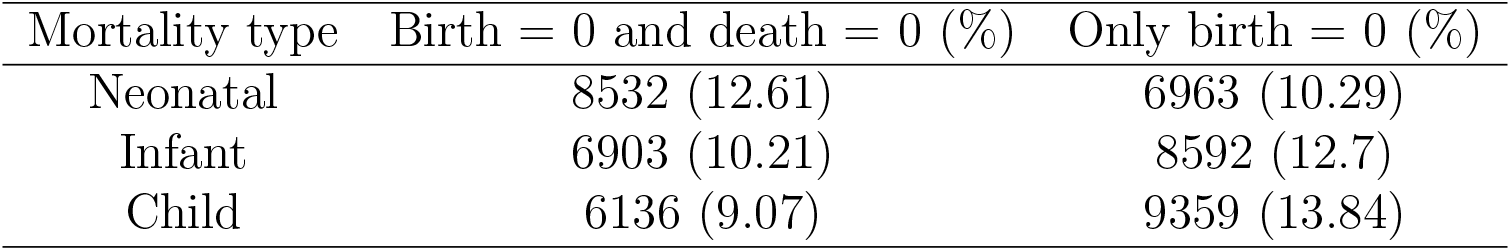
Counts and percentage of 0 birth in municipality-year level for three types of mortality.

**Figure 10.**
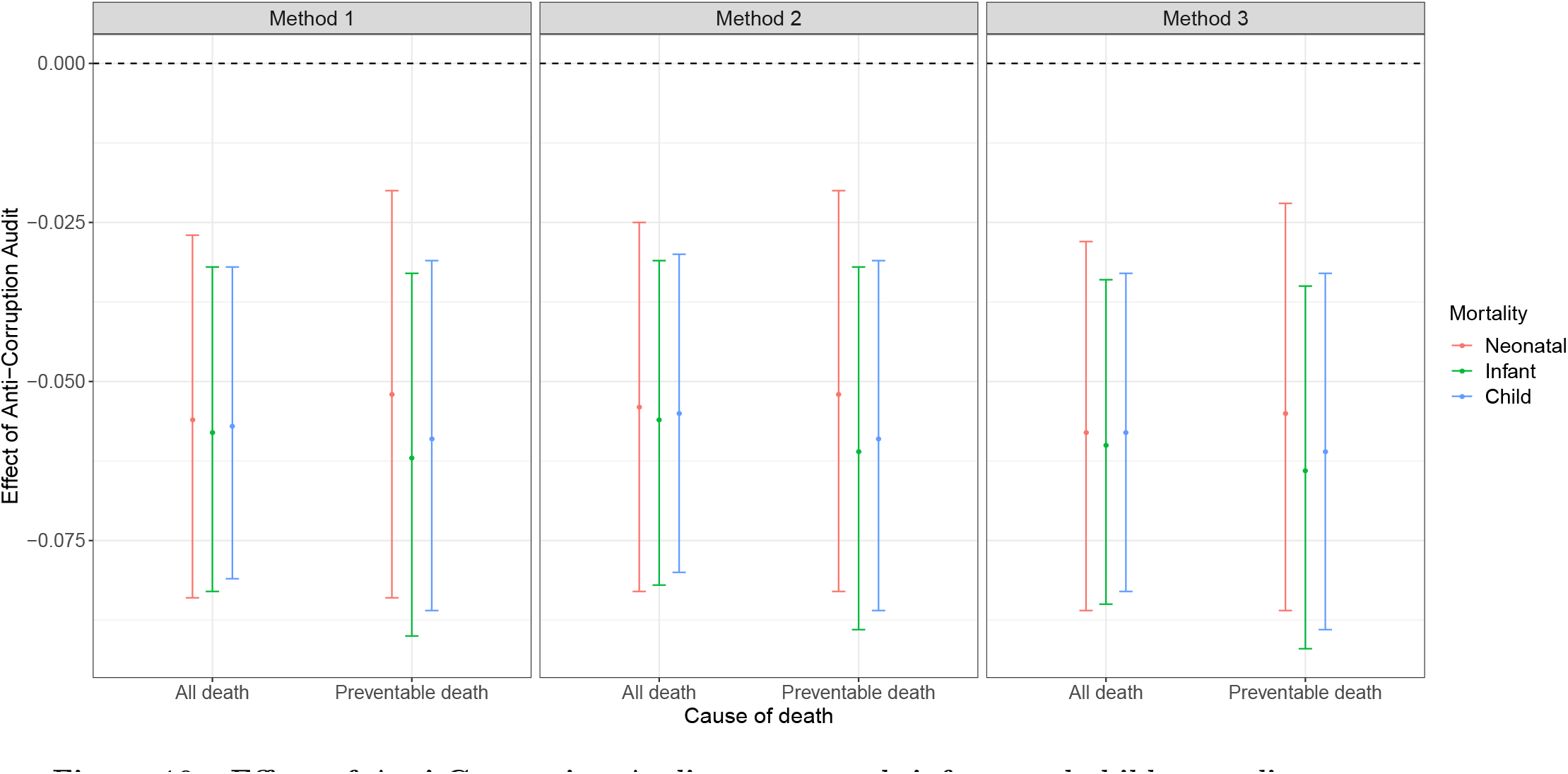
Effect of Anti-Corruption Audits on neontal, infant, and child mortality, by cause of death, using three methods to deal with the offset. The dots are pi.

Given the use of Poisson models, results reported above reflect relative mortality reductions. The impact of anti-corruption audits is further clarified when considering the absolute number of lives saved by the intervention. Altogether, municipalities randomly selected for corruption audits had XX fewer children perish by their fifth birthday than the counterfactual in which they had not been audited. Included in this decline in child mortality are XX fewer neonatal deaths (ages 0 to 28 days) and XX fewer post-neonatal deaths (ages 29 days to 1 year). The intervention disproportionately saved lives of nonwhite Brazilians. This population represents half (CHECK) of all births in Brazil, but about two-thirds (CHECK) of children saved by the intervention. More specifically, nonwhite Brazilians had XX fewer children perish (vs. XX for white Brazilians), including XX fewer neonatal deaths and XX post-neonatal deaths (vs. XX and XX, respectively). This disproportionate impact not only reflects the intervention’s greater relative effects on non-white Brazilians (see Figure X), but also existing racial disparities in which non-white Brazilians suffer from higher absolute mortality rates (see Table 1).

## Appendix C: Additional Tables

**Table 6.**
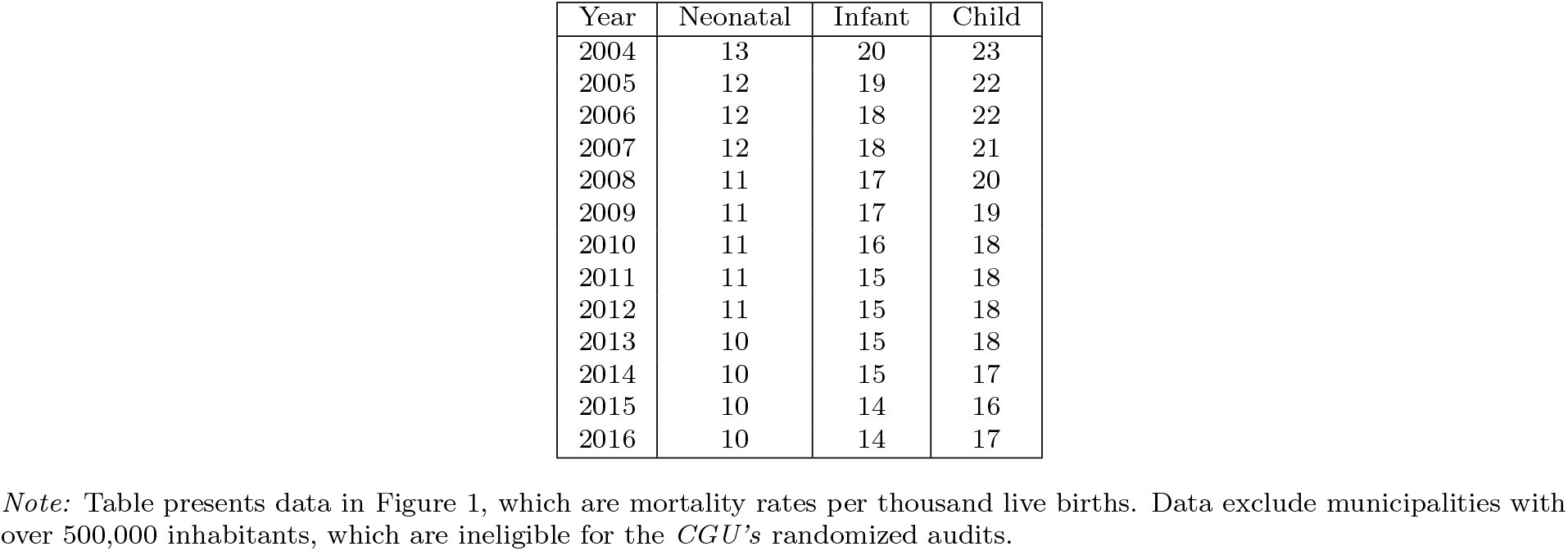
Early Life Mortality by Year (2004-2016)

**Table 7.** Effect of Anti-Corruption Audits on Early-Life Mortality.

| Cause | Age | All Races | White | Nonwhite |
| --- | --- | --- | --- | --- |
| All Deaths | Neonatal | -0.067 (-0.083, -0.05) | -0.033 (-0.061, -0.005) | -0.081 (-0.103, -0.057) |
| All Deaths | Infant | -0.073 (-0.086, -0.059) | -0.036 (-0.06, -0.013) | -0.082 (-0.101, -0.063) |
| All Deaths | Child | -0.073 (-0.085, -0.06) | -0.04 (-0.061, -0.018) | -0.082 (-0.099, -0.064) |
| Preventable Deaths | Neonatal | -0.065 (-0.084, -0.046) | -0.033 (-0.065, 0.001) | -0.081 (-0.107, -0.055) |
| Preventable Deaths | Infant | -0.079 (-0.095, -0.063) | -0.038 (-0.065, -0.008) | -0.095 (-0.118, -0.072) |
| Preventable Deaths | Child | -0.077 (-0.092, -0.061) | -0.035 (-0.062, -0.008) | -0.092 (-0.112, -0.071) |

*Note:* Table presents results in Figure 2, which shows the effect of audits on neonatal, infant and child mortality (by race). Point estimates are from Bayesian Hierarchical Poisson regressions, with 95% credible intervals in parentheses. Audit effects are expressed in terms of relative mortality reduction. Results are from Model 1 and are fitted with data for all deaths and preventable causes of death (respectively). Specifications are shown fitting the model using data for all races, whites, and nonwhites (respectively).

**Table 8.** Temporal Effects of Anti-Corruption Audits on Early-Life Mortality.

| Years | Race | Neonatal | Infant | Child |
| --- | --- | --- | --- | --- |
| 1 | All Races | -0.017 (-0.041, 0.007) | -0.026 (-0.045, -0.006) | -0.025 (-0.043, -0.007) |
| 2 | All Races | -0.043 (-0.067, -0.019) | -0.052 (-0.071, -0.032) | -0.051 (-0.069, -0.032) |
| 3 | All Races | -0.033 (-0.059, -0.007) | -0.039 (-0.059, -0.019) | -0.041 (-0.06, -0.022) |
| 4 | All Races | -0.062 (-0.088, -0.036) | -0.068 (-0.089, -0.047) | -0.066 (-0.085, -0.047) |
| 5 | All Races | -0.078 (-0.103, -0.051) | -0.083 (-0.104, -0.061) | -0.08 (-0.099, -0.06) |
| 6 | All Races | -0.088 (-0.114, -0.061) | -0.094 (-0.116, -0.071) | -0.096 (-0.116, -0.076) |
| 7 | All Races | -0.117 (-0.145, -0.089) | -0.125 (-0.148, -0.102) | -0.115 (-0.136, -0.093) |
| 8 | All Races | -0.155 (-0.183, -0.127) | -0.15 (-0.173, -0.126) | -0.142 (-0.163, -0.119) |
| 9 | All Races | -0.141 (-0.171, -0.11) | -0.146 (-0.171, -0.121) | -0.138 (-0.161, -0.114) |
| 10 | All Races | -0.133 (-0.166, -0.099) | -0.15 (-0.177, -0.123) | -0.141 (-0.166, -0.116) |
| 1 | Whites | 0.044 (0.004, 0.086) | 0.019 (-0.014, 0.053) | 0.01 (-0.019, 0.042) |
| 2 | Whites | 0.002 (-0.04, 0.046) | 0.001 (-0.033, 0.036) | 0.01 (-0.032, 0.033) |
| 3 | Whites | 0.024 (-0.019, 0.07) | 0.027 (-0.009, 0.064) | 0.025 (-0.008, 0.06) |
| 4 | Whites | -0.027 (-0.07, 0.02) | -0.016 (-0.052, 0.021) | -0.024 (-0.057, 0.01) |
| 5 | Whites | -0.068 (-0.112, -0.022) | -0.07 (-0.104, -0.034) | -0.069 (-0.102, -0.035) |
| 6 | Whites | -0.029 (-0.077, 0.02) | -0.038 (-0.077, 0.002) | -0.046 (-0.08, -0.011) |
| 7 | Whites | -0.079 (-0.127, -0.028) | -0.078 (-0.117, -0.037) | -0.07 (-0.106, -0.033) |
| 8 | Whites | -0.089 (-0.138, -0.037) | -0.076 (-0.117, -0.035) | -0.07 (-0.108, -0.03) |
| 9 | Whites | -0.058 (-0.114, 0) | -0.068 (-0.112, -0.021) | -0.068 (-0.109, -0.025) |
| 10 | Whites | -0.027 (-0.087, 0.037) | -0.028 (-0.077, 0.023) | -0.029 (-0.074, 0.018) |
| 1 | Nonwhites | -0.046 (-0.079, -0.013) | -0.044 (-0.071, -0.017) | -0.037 (-0.062, -0.012) |
| 2 | Nonwhites | -0.071 (-0.104, -0.036) | -0.073 (-0.1, -0.045) | -0.071 (-0.096, -0.045) |
| 3 | Nonwhites | -0.041 (-0.076, -0.006) | -0.055 (-0.083, -0.027) | -0.058 (-0.083, -0.032) |
| 4 | Nonwhites | -0.08 (-0.115, -0.044) | -0.094 (-0.122, -0.065) | -0.088 (-0.113, -0.061) |
| 5 | Nonwhites | -0.078 (-0.114, -0.04) | -0.079 (-0.109, -0.047) | -0.079 (-0.106, -0.051) |
| 6 | Nonwhites | -0.096 (-0.132, -0.057) | -0.094 (-0.126, -0.063) | -0.095 (-0.123, -0.066) |
| 7 | Nonwhites | -0.134 (-0.172, -0.095) | -0.137 (-0.169, -0.104) | -0.122 (-0.151, -0.092) |
| 8 | Nonwhites | -0.192 (-0.229, -0.153) | -0.182 (-0.214, -0.148) | -0.175 (-0.205, -0.144) |
| 9 | Nonwhites | -0.186 (-0.227, -0.144) | -0.18 (-0.214, -0.145) | -0.169 (-0.2, -0.136) |
| 10 | Nonwhites | -0.188 (-0.231, -0.142) | -0.197 (-0.233, -0.159) | -0.185 (-0.218, -0.15) |

*Note:* Table presents results in Figure 3, which shows the temporal effect of audits on neonatal, infant and child mortality (by race). “Years” refer to years after anti-corruption audit. Point estimates are from Bayesian Hierarchical Poisson regressions, with 95% credible intervals in parentheses. Audit effects are expressed in terms of relative mortality reduction. Specifications employ Model 2, fitted with data from all races, whites and nonwhites.

**Table 9.**
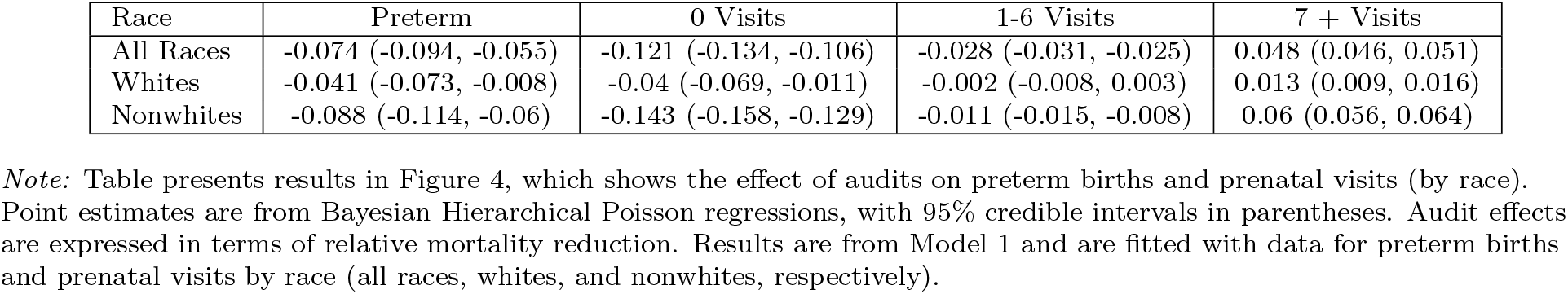
Effect of Anti-Corruption Audits on Preterm Births and Prenatal Visits.

| Race | Preterm | 0 Visits | 1-6 Visits | 7 + Visits |
| --- | --- | --- | --- | --- |
| All Races | -0.074 (-0.094, -0.055) | -0.121 (-0.134, -0.106) | -0.028 (-0.031, -0.025) | 0.048 (0.046, 0.051) |
| Whites | -0.041 (-0.073, -0.008) | -0.04 (-0.069, -0.011) | -0.002 (-0.008, 0.003) | 0.013 (0.009, 0.016) |
| Nonwhites | -0.088 (-0.114, -0.06) | -0.143 (-0.158, -0.129) | -0.011 (-0.015, -0.008) | 0.06 (0.056, 0.064) |

*Note:* Table presents results in Figure 4, which shows the effect of audits on preterm births and prenatal visits (by race). Point estimates are from Bayesian Hierarchical Poisson regressions, with 95% credible intervals in parentheses. Audit effects are expressed in terms of relative mortality reduction. Results are from Model 1 and are fitted with data for preterm births and prenatal visits by race (all races, whites, and nonwhites, respectively).

**Table 10.** Temporal Effects of Anti-Corruption Audits on Preterm Births.

| Years | All Races | White | Nonwhite |
| --- | --- | --- | --- |
| 1 | -0.011 (-0.038, 0.018) | 0.021 (-0.028, 0.071) | -0.03 (-0.069, 0.011) |
| 2 | -0.046 (-0.075, -0.016) | 0.001 (-0.049, 0.053) | -0.086 (-0.126, -0.045) |
| 3 | -0.061 (-0.091, -0.032) | -0.003 (-0.056, 0.051) | -0.088 (-0.129, -0.046) |
| 4 | -0.084 (-0.114, -0.053) | -0.052 (-0.102, 0) | -0.093 (-0.135, -0.049) |
| 5 | -0.065 (-0.096, -0.034) | -0.034 (-0.085, 0.02) | -0.057 (-0.102, -0.012) |
| 6 | -0.103 (-0.133, -0.071) | -0.05 (-0.101, 0.006) | -0.122 (-0.165, -0.077) |
| 7 | -0.131 (-0.163, -0.097) | -0.095 (-0.147, -0.04) | -0.134 (-0.181, -0.088) |
| 8 | -0.163 (-0.196, -0.13) | -0.112 (-0.167, -0.053) | -0.199 (-0.244, -0.152) |
| 9 | -0.152 (-0.187, -0.115) | -0.09 (-0.15, -0.024) | -0.195 (-0.241, -0.146) |
| 10 | -0.162 (-0.199, -0.123) | -0.078 (-0.144, -0.01) | -0.213 (-0.265, -0.16) |

*Note:* Table presents results in Figure 5, which shows the temporal effect of audits on prenatal births (by race). “Years” refer to years after anti-corruption audit. Point estimates are from Bayesian Hierarchical Poisson regressions, with 95% credible intervals in parentheses. Audit effects are expressed in terms of relative reduction. Specifications employ Model 2, fitted with data from all races, whites and nonwhites.

## Footnotes

1 Moreover, Zamboni & Litschig (2018) find that temporarily increasing a municipality’s audit risk does not affect health worker absenteeism (according to user satisfaction surveys).^42^

